# Zero warm ischemia time in whole-eye transplantation through retrograde extracorporeal perfusion

**DOI:** 10.1101/2025.10.20.25338319

**Authors:** Anthony C. Wang, Yasamin Sadat Mousavi Motlagh, Iden Amiri, H. Milan Samarage, Warwick J. Peacock, Aya Barzelay Wollman

**Affiliations:** Department of Neurosurgery, University of California, Los Angeles, Los Angeles, CA 90095; Stein Eye Institute, Los Angeles, CA 90095; Department of Ophthalmology, University of North Carolina at Chapel Hill, Chapel Hill, NC 27517; David Geffen School of Medicine, University of California, Los Angeles, Los Angeles, CA 90095

**Author notes:** Corresponding Author: Aya Barzelay-Wollman, MD, PhD, Department of Ophthalmology, University of California, Los Angeles, 100 Stein Plaza Driveway, Los Angeles, CA 90095. Funding: Advanced Research Projects Agency for Health (ARPA-H).

**Keywords:** whole-eye transplantation, warm ischemia time, retrograde arterial perfusion, optic nerve coaptation, ocular allotransplantation

## Abstract

**Purpose:** To develop and demonstrate the anatomical feasibility of a human whole-eye transplantation (WET) protocol that eliminates warm ischemia time (WIT) through continuous retrograde arterial perfusion, enabling uninterrupted ocular oxygenation throughout the transplantation process.

**Design:** Experimental cadaveric feasibility study.

**Subjects:** Two fresh human cadaveric specimens used to simulate a complete donor-to-recipient WET sequence.

**Methods:** A modified fronto-temporo-orbito-zygomatic craniotomy with anterior clinoidectomy was performed on each donor to expose the anterior cerebral artery (ACA), middle cerebral artery (MCA), and ophthalmic artery (OA). The ACA was cannulated to permit continuous retrograde perfusion of the ophthalmic circulation while the globe, with preserved optic nerve and extraocular muscles, was harvested en bloc. Recipient preparation involved mobilization of the internal maxillary artery (IMAX) in the infratemporal fossa and removal of the native globe. Orthotopic transplantation was achieved with MCA–IMAX and superior ophthalmic vein (SOV– SOV) microvascular anastomoses and optic nerve coaptation under microscopy, maintaining continuous simulated perfusion through the ACA cannula.

**Main Outcome Measures:** Anatomical feasibility of achieving zero warm ischemia time during the entire transplantation sequence, confirmed by optical coherence tomography (OCT), intraoperative microscopy, and photographic documentation of vascular and neural integration.

**Results:** Continuous retrograde perfusion was maintained throughout donor harvest and transplantation, achieving zero WIT across both specimens. OCT verified luminal patency and vessel continuity after ACA cannulation and MCA–IMAX anastomosis. The IMAX provided a robust, tension-free arterial recipient site with consistent caliber and orientation. Optic nerve coaptation and extraocular muscle reattachment were anatomically aligned, with sequential imaging confirming vascular and neural integration. The technique proved reproducible across both dissections, supporting its feasibility for perfused, anatomically integrated ocular allotransplantation.

**Conclusions:** This study establishes a reproducible human WET protocol achieving zero WIT via continuous retrograde extracorporeal perfusion. The approach provides a foundational anatomical framework for future functional and preclinical studies aiming to preserve retinal viability and enable neurovascularly integrated whole-eye transplantation.

## INTRODUCTION

Whole eye transplantation (WET) represents the next frontier in facial composite allotransplantation, involving the *en bloc* transfer of ocular, vascular, and neural components. Although advances have been made in partial orbital and facial transplantation, complete restoration of a perfused, anatomically integrated eye with potential for visual recovery remains an elusive goal.^1–3^ The first reported human case of WET involved harvesting the globe with an attached optic nerve and vascular pedicle, followed by microvascular anastomosis to the recipient’s superficial temporal artery (STA) and vein, with reattachment of extraocular muscles. Histopathology demonstrated Wallerian degeneration of the optic nerve without evidence of functional coaptation. The result was a viable, vascularized globe with preserved structural integrity but no light perception one year post-operatively.^4,5^

One of the principal challenges in allotransplantation lies in mitigating ischemic injury during both procurement and transplantation. In the broader field of organ transplantation, ischemia is typically divided into cold ischemia time (CIT), the duration the organ is stored at hypothermic conditions and warm ischemia time (WIT), the time the organ remains at body temperature without blood supply.^6,7^ Warm ischemia is particularly harmful due to sustained metabolic activity in the absence of oxygenation.^8,9^ The retina is exceptionally susceptible to ischemic insult, and even carefully optimized surgical WIT may not be sufficient to preserve function^10,11^, given the retina’s high metabolic demands.^12^ Techniques designed to minimize the WIT are a cornerstone of successful organ transplantation. In the case referenced above, CIT was reported as 2 hours and 59 minutes, while WIT was described as approximately 25 minutes until arterial reperfusion was achieved; however, the precise timing of complete arterial and venous flow restoration remains uncertain.^4,5^ There is a critical need to minimize ischemia time and, ideally, enable continuous retinal oxygenation during the entire transplant procedure.

To overcome this limitation, we performed a feasibility study to demonstrate a novel WET protocol developed to reduce WIT to zero by incorporating retrograde arterial cannulation and continuous perfusion, modeled after extracorporeal circulation systems such as extracorporeal membrane oxygenation (ECMO). In parallel with this objective, researchers at the Bascom Palmer Eye Institute and the University of Miami Miller School of Medicine have engineered a portable eye-ECMO system that maintains oxygenated flow through the donor eye *ex vivo* using a custom cannula and eye holder platform. Initial institutional reports describe its successful use in preserving retinal viability for several hours post-recovery, marking a critical step toward perfusion-based eye preservation strategies in WET.^13^

## METHODS

Two fresh human cadavers were utilized to simulate a full donor-recipient transplantation sequence. Procedures were performed under institutional anatomic research protocols. The specimens were placed in the supine position and prepared for craniofacial exposure. Representative stepwise procedural images are presented in the results section (Figures 1–5).

**Figure 1.**
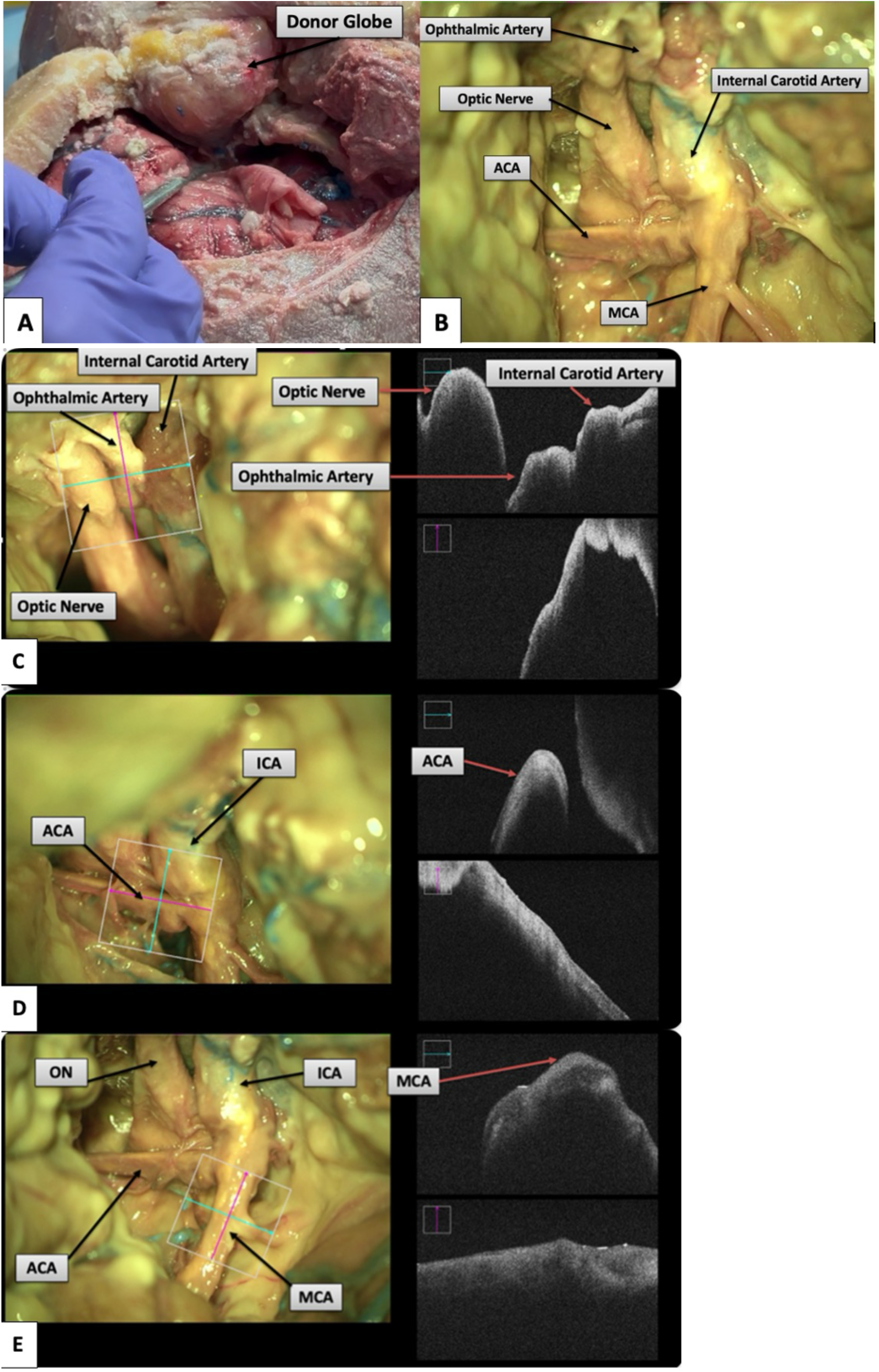
Donor-side exposure. (A) Modified fronto-temporo-orbito-zygomatic craniotomy with anterior clinoidectomy provides adequate exposure for donor globe harvest. (B) Microscopic view of the paraclinoid and supraclinoid segments of the ICA, and the optic nerve. OCT demonstrating patency of the ICA and OA. (D) OCT demonstrating patency of the ACA, suitable for eye-ECMO cannulation. (E) OCT demonstrating patency of the MCA, suitable for microvascular anastomosis.

**Figure 2.**
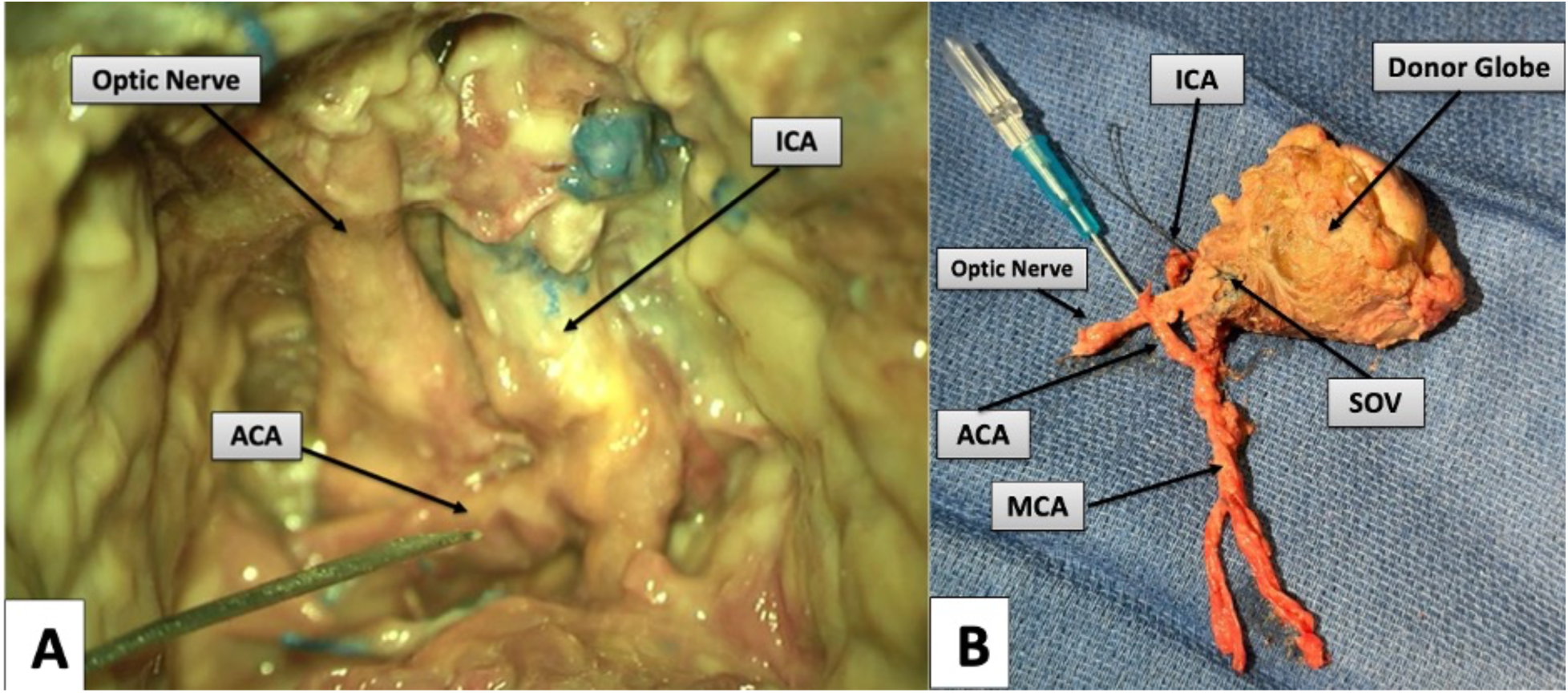
ACA cannulation and donor globe isolation. (A) Cannulation of the ACA (here, with a 24-gauge peripheral venous access catheter) and suitable for continuous perfusion. (B) Isolated donor globe with intact and cannulated arterial pedicle (ACA, MCA, OA), preserved venous outlet channel (SOV), and entire length of the optic nerve.

**Figure 3.**
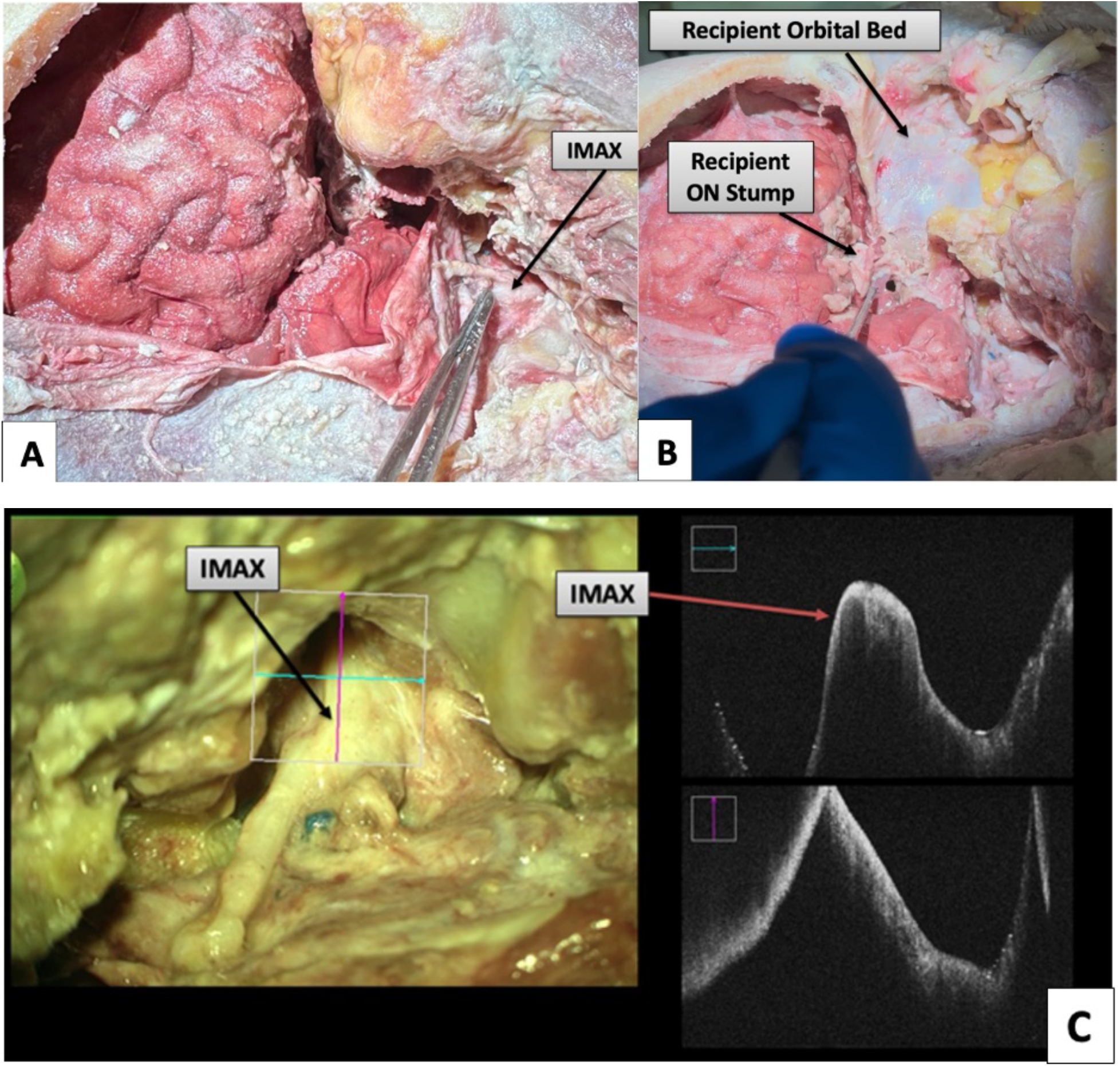
Recipient-side exposure. (A) Full fronto-temporo-orbito-zygomatic approach exposing the IMAX in the infratemporal fossa. (B) Post-enucleation orbital cavity after anterior clinoidectomy, with preserved recipient optic nerve (ON) stump. (C) Microscopic view of IMAX after mobilization, and OCT demonstrating a patent lumen and intact vessel wall, with a caliber well-suited for MCA anastomosis.

**Figure 4.**
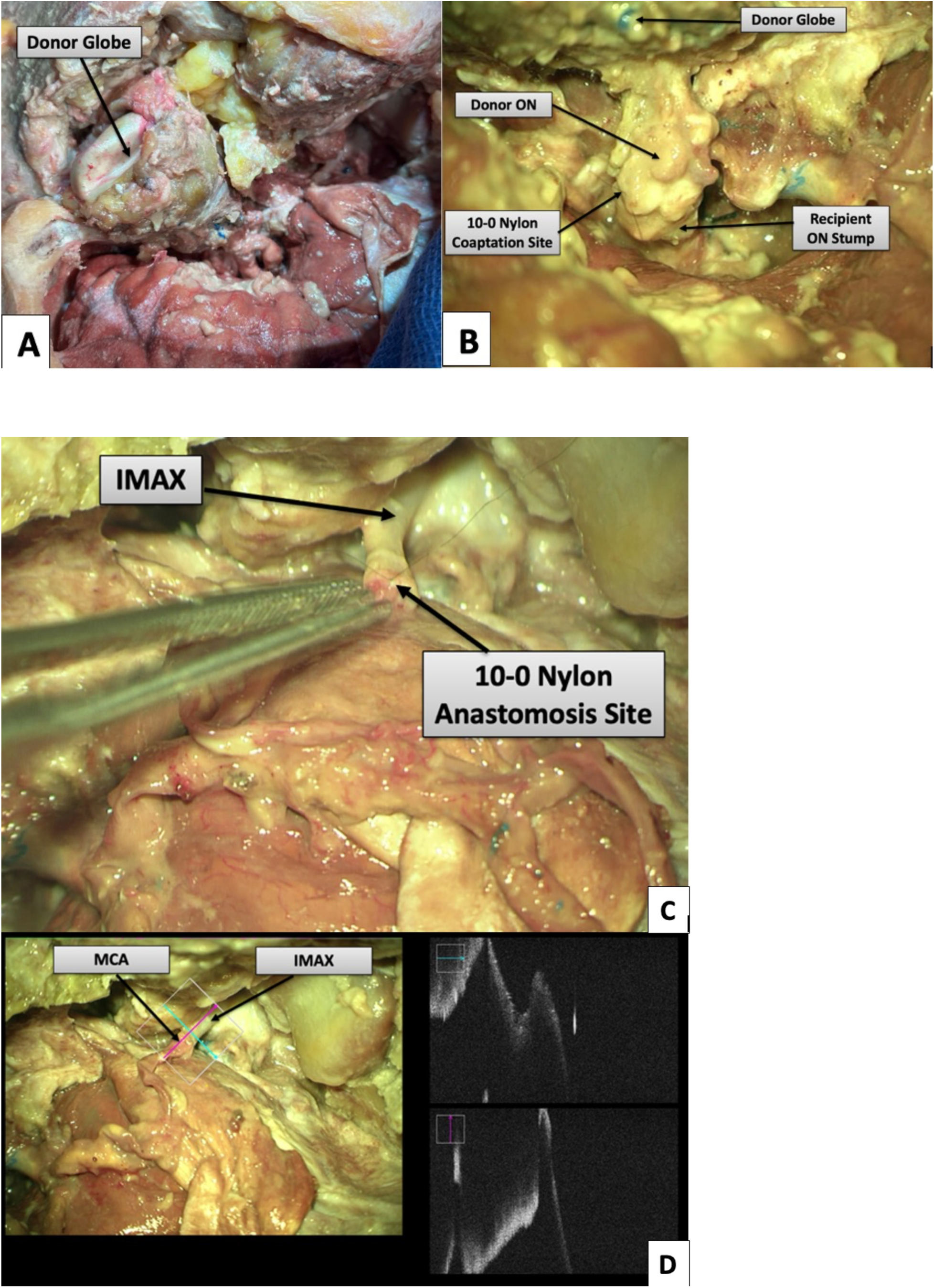
Orthotopic donor globe placement and neurovascular integration. (A) Donor globe positioned within the recipient orbit with vascular pedicle aligned toward the recipient IMAX. (B) Optic nerve coaptation between donor and recipient stumps using interrupted 10-0 nylon sutures. (C) MCA–IMAX prior to completion of end-to-end microvascular anastomosis. OCT of the completed anastomosis demonstrating continuity of the lumen.

**Figure 5.**
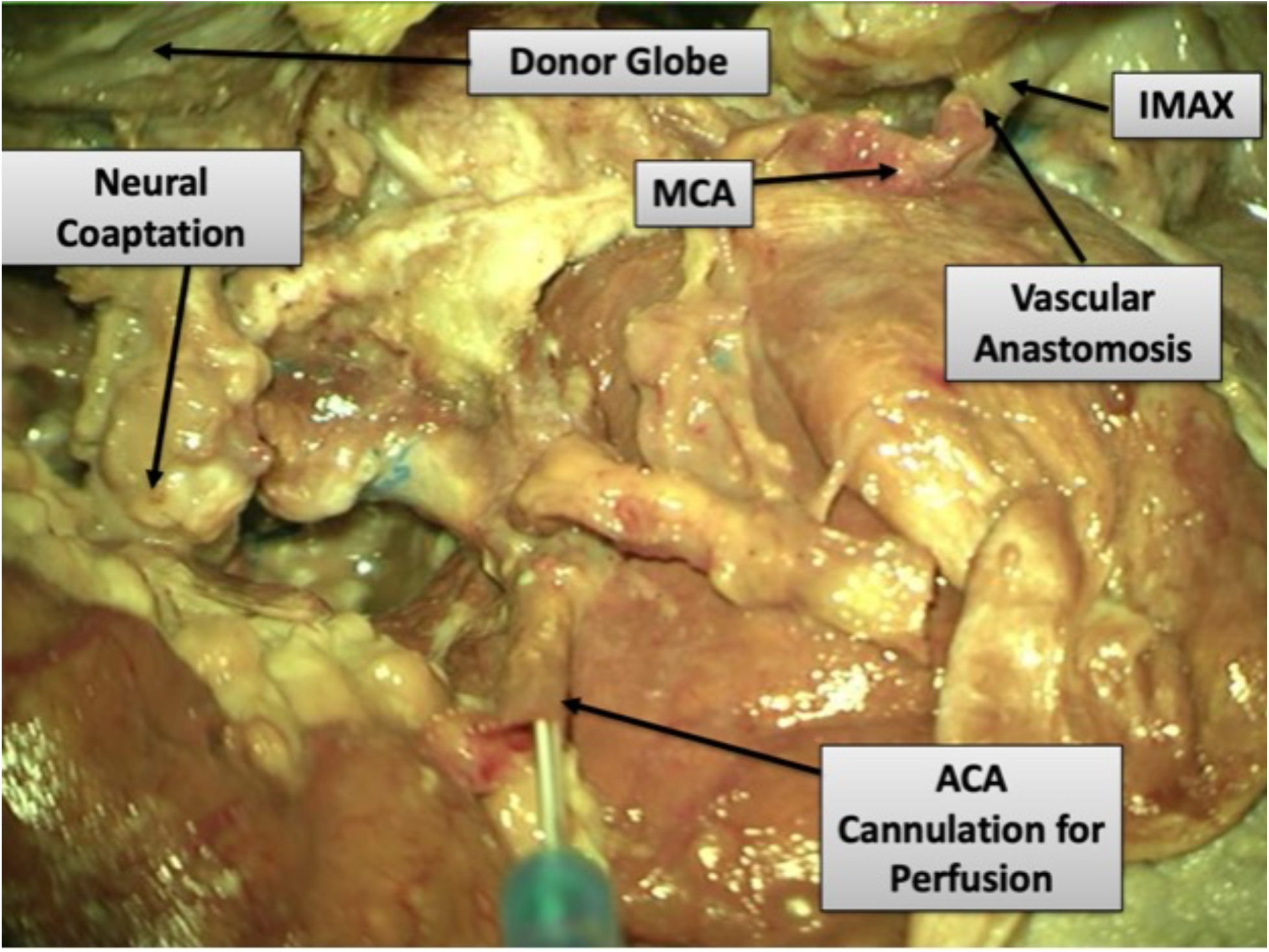
Final configuration after transplantation. Microscopic overview of the final construct demonstrating donor MCA to recipient IMAX anastomosis, ACA still cannulated for simulated continuous perfusion, and optic nerve coapted to recipient stump.

### Donor Procedure

A modified two-piece fronto-temporo-orbito-zygomatic craniotomy^14^ is performed with anterior clinoidectomy to provide a wide access corridor to the optic canal, the superior orbital fissure, the cavernous sinus, the middle cerebral artery (MCA), and the anterior cerebral artery (ACA). Using standard microsurgical techniques, the optic chiasm and the full extent of the A1 segment of the ACA and the M1 and proximal M2 segments of the MCA are exposed (see Figure 1A–F, Results). The distal dural ring is incised, and the cavernous segment of the internal carotid artery (ICA) is exposed below the take-off of the ophthalmic artery (OA). The oculomotor nerve, trochlear nerve, and abducens nerve, along with their innervated extraocular muscles, and the superior ophthalmic vein (SOV) are then identified, following the SOV into the cavernous sinus.

Arterial cannulation is performed at this point and can be achieved through either an M2 branch of the MCA or through the A1 segment of the ACA. In this study, a robust ACA allowed for cannulation of the ACA (see Figure 2A–B, Results). The ICA proximal to the take-off of the OA is permanently ligated and the proximal M1 segment of the MCA is temporarily occluded, allowing for retrograde perfusion of the globe via the OA. The optic nerve is divided at the optic chiasm, SOV is divided at the cavernous sinus, and the globe is removed en bloc, being careful to ligate and divide the inferior orbital vein sharply, long segments of the extraocular muscle bundles are preserved for attachment to recipient extraocular musculature, along with adjacent periorbital soft tissues.

### Recipient Procedure

A standard fronto-temporo-orbito-zygomatic craniotomy^15^ is performed fully exposing the anterior and middle cranial fossae while providing wide exposure to the orbit and the infratemporal fossa. The internal maxillary artery (IMAX) is commonly used as a high-flow donor vessel in cerebral revascularization and provides a robust and consistent arterial pedicle that is well-suited in size. IMAX is then harvested in the infratemporal fossa; however, in order to obtain additional length, we have also performed donor artery procurement via an endoscope-assisted technique.^16^ The recipient’s native globe is then removed, taking care to section the optic nerve, SOV, and extraocular muscles distally (see Figure 3A–C, Results).

The donor globe is then positioned in the orbital cavity, and optic nerve coaptation is performed, placing two perineural sutures. The recipient IMAX is anastomosed to the donor eye MCA in an end-to-end fashion, and SOV-to-SOV anastomosis is performed before ligating the inferior orbital vein. The ACA is then de-cannulated, back-bled, and ligated. In this manner, perfusion is maintained to the donor eye throughout, with no WIT (see Figure 4A–D, Results). Recipient extraocular muscles are then sutured to donor muscle tendons.

In the final configuration, the donor MCA–IMAX anastomosis was intact, ACA cannulation allowed simulated continuous perfusion, and the optic nerve coaptation was anatomically aligned (Figure 5). Sequential photographs and schematic overlays illustrate each major step from the donor-side exposure to the final placement and neurovascular integration. These results demonstrate the anatomical feasibility of simultaneous vascular and neural integration using this skull base–assisted model.

## RESULTS

Across two fresh human cadaveric specimens, we simulated the complete donor-to-recipient whole-eye transplantation (WET) workflow and documented each step with intraoperative microscopy and optical coherence tomography (OCT). Representative images are shown in Figures 1–5.

### Donor exposure and arterial access

A modified fronto-temporo-orbito-zygomatic craniotomy with anterior clinoidectomy provided a wide skull-base corridor to the optic canal, superior orbital fissure, cavernous sinus, middle cerebral artery (MCA), and anterior cerebral artery (ACA). The optic chiasm, A1 segment of the ACA, and M1/proximal M2 segments of the MCA were exposed; the distal dural ring was incised and the cavernous internal carotid artery (ICA) identified below the ophthalmic artery (OA) origin. The superior ophthalmic vein (SOV) and cranial nerves III, IV, and VI were localized by tracking the SOV into the cavernous sinus. OCT confirmed luminal patency of ICA/OA, ACA, and MCA (Figure 1C–E).

### Cannulation and donor harvest

Arterial cannulation was performed at the skull base; although feasible via an M2 branch, in this study the robust ACA allowed ACA cannulation (Figure 2A; 24-gauge catheter). The ICA proximal to the OA take-off was permanently ligated and the proximal M1 was temporarily occluded to create retrograde perfusion to the globe via the OA. The optic nerve was divided at the chiasm, the SOV at the cavernous sinus, and the globe removed en bloc with preserved long extraocular muscle segments and periorbital soft tissues; the inferior orbital vein was ligated. The isolated donor specimen retained an intact cannulated arterial pedicle (ACA–MCA–OA) and preserved venous outlet (SOV) (Figure 2B).

### Recipient preparation

A standard fronto-temporo-orbito-zygomatic craniotomy exposed the anterior/middle cranial fossae, orbit, and infratemporal fossa. The internal maxillary artery (IMAX) was mobilized in the infratemporal fossa (with endoscope-assisted harvest used when additional length was needed). The native globe was removed with distal sectioning of the optic nerve, SOV, and extraocular muscles. OCT confirmed IMAX lumen integrity and vessel wall continuity suitable for microvascular anastomosis (Figure 3).

### Orthotopic placement, anastomoses, and perfusion management

The donor globe was placed orthotopically; optic nerve coaptation was achieved with two perineural sutures (Figure 4B, interrupted 10-0 nylon). An end-to-end MCA–IMAX arterial anastomosis and SOV-to-SOV venous anastomosis were completed under microscopy, while simulated perfusion was maintained continuously through the ACA cannula, achieving zero warm ischemia time (WIT). After establishing inflow, the ACA was de-cannulated, back-bled, and ligated, with uninterrupted perfusion preserved through the definitive arterial anastomosis. OCT demonstrated lumen continuity of the completed anastomosis (Figure 4D).

### Extraocular muscles and construct integrity

Recipient extraocular muscles were sutured to donor muscle tendons. Final microscopy confirmed an anatomically aligned optic nerve coaptation and an intact donor MCA-to-recipient IMAX anastomosis; at the documentation stage, the ACA remained cannulated to demonstrate ongoing simulated perfusion (Figure 5). Sequential intraoperative photographs and schematic overlays verified each step and the final neurovascular integration.

### Overall outcome

Across both specimens, continuous simulated perfusion was maintained throughout procurement and transplantation with no WIT, vascular patency was corroborated by OCT before and after anastomoses, and the final construct demonstrated reproducible combined vascular and neural integration suitable for subsequent functional studies.

## DISCUSSION

The retina is among the most metabolically demanding tissues in the body, and its photoreceptors are particularly vulnerable to ischemia. Following cessation of circulation, the retina undergoes a rapid decline in function due to its exceptionally high metabolic requirements.^12,17^ Experimental evidence from *ex vivo* primate and human models has demonstrated that detectable electroretinographic responses may persist for only minutes after ischemia onset, with photoreceptor and ON-bipolar cell light responses being recoverable when eyes are re-perfused within approximately 15–20 minutes post-mortem.^10,18,19^ Beyond this interval, functional recovery declines precipitously.^10^ Classic morphologic and electrophysiologic studies similarly show that inner retinal neurons (e.g., ganglion cells) swell within minutes of complete ischemia, while early outer segment disruption in photoreceptors emerges within the first hour, indicating that even brief warm ischemia can precipitate irreversible injury.^18,19^ Photoreceptor degeneration begins within 12 hours after onset of ischemia, with marked thinning of the outer nuclear layer by day 3 and substantial structural loss by day 7.^18^ Other models of transient retinal ischemia have demonstrated significant outer retinal degeneration within the first 24 hours, with more than 50% photoreceptor cell loss by day 3.^20,21^ These observations reinforce the central premise of this protocol: initiating ocular perfusion as early as possible is essential to preserving retinal viability.

In other fields of transplantation, continuous or near-continuous perfusion has been successfully applied to extend graft viability and reduce ischemia–reperfusion injury. In liver transplantation, techniques such as hypothermic oxygenated perfusion (HOPE) and normothermic machine perfusion (NMP) have been shown to maintain cellular metabolism, reduce injury, and improve post-transplant function compared to static cold storage.^21,22^ A recent multicenter randomized controlled trial in the United States further demonstrated that normothermic machine perfusion of donor livers can improve preservation quality and enable functional assessment before implantation compared with conventional cold storage.^22^ Adapting these principles to ocular transplantation, we propose to cannulate the ACA for eye-ECMO perfusion prior to ligation of the native arterial supply through the OA, thus enabling zero WIT. This technique is feasible for immediate integration with portable perfusion systems, such as those developed for other vascularized composite allografts.^23,24^ Collaborating with biomedical engineering teams, such as the group at the University of Miami that has already reported development of a portable eye-ECMO system capable of maintaining oxygenated flow through an enucleated donor eye^13,25,26^, may enable real-time retinal perfusion throughout procurement and transfer. This approach has the potential to preserve retinal viability in a way that has not been possible in other proposed human WET procedures.^4,27^

A number of cadaveric studies have evaluated the technical feasibility of whole face and eye transplantation, as summarized in recent systematic reviews^1–3^, but there are no published technique descriptions of WET alone in humans.^4,5^ Our described technique differs dramatically from that performed by the NYU group, however. In our technique, the donor eye is perfused retrograde through the MCA, which allows for a larger arterial anastomosis than the previously-described STA-OA anastomosis, and provides greater length of arterial pedicle. Critically, this allows us to initiate extracorporeal perfusion without WIT via the ACA, and to continue this perfusion during arterial microvascular anastomosis, maintaining continuous simulated perfusion that targets zero WIT.

Another advantageous feature of our technique is the use of the IMAX as the arterial supply to the donor eye. The IMAX, a terminal branch of the external carotid artery, demonstrates a relatively consistent caliber, reported average diameters around 2.0 mm with relatively low inter-individual variability and a predictable anatomic course relative to the lateral pterygoid muscle.^28,29^ A long segment can be harvested with endoscopic assistance^16^, and when paired with an MCA recipient artery, provides adequate arterial length. By contrast, the STA exhibits substantial variability in diameter, branching pattern, and distant location to the orbit, as well as significantly lower flow rates.^30^ Use of the IMAX therefore increases arterial flow, improves reproducibility, and facilitates a tension-free anastomosis.^31–33^ Practically, this yields a more reproducible, caliber-matched, and straight recipient segment for end-to-end suturing to donor M1 than is typically achievable with STA, thereby simplifying orientation and minimizing anastomotic tension.

This study has several limitations. Being cadaveric in nature, it does not permit functional assessment of retinal perfusion or vision recovery, and the heparinized saline flush that we used cannot replicate the physiologic oxygen and nutrient delivery of live circulation. In addition, we did not assess the adequacy of venous outflow via SOV-SOV anastomosis. Future work should incorporate live animal models and *ex vivo* perfusion experiments to measure retinal oxygenation, flow rates, and structural preservation.

Additionally, although retinal and graft survival have been achieved in animal models, including cold-blooded vertebrates and select mammalian experiments, significant technical barriers remain, especially in translating these findings to human clinical application.^34^ Furthermore, clinician–scientist perspectives emphasize that overcoming the obstacles related to ocular perfusion, neural regeneration, and immune modulation is essential for advancing WET from an experimental concept to a viable clinical reality.^24^

Adaptation of a standardized portable perfusion platform for ocular transplantation similar to the Organ Care System, a clinically validated portable normothermic perfusion device now used for thoracic and abdominal organ transplantation^23^, will be essential for translating this anatomical protocol into a viable clinical procedure.

## CONCLUSION

Through these cadaveric dissections, we demonstrate the technical feasibility of the complete WET workflow. By employing standard skull base approaches, microvascular neurosurgical techniques, and the portable eye-ECMO system, we describe a WET protocol that allows for continuous perfusion with no ischemia time, robust arterial supply and venous drainage, and the ability to attach extraocular muscles directly to existing innervated musculature. The vascular and neural anastomosis sites were clearly delineated under microscopy. This foundational platform represents a significant technical advancement from previously reported WET techniques, eliminating WIT. Integration with portable perfusion systems and translation to live experimental models will be essential next steps toward successfully orchestrating the first functional human whole-eye transplant.

## Data Availability

All data produced in the present work are available upon reasonable request to the authors.

## Acknowledgements

The authors wish to thank individuals who donate their bodies and tissues for the advancement of education and research.

## AI Disclosure

The authors did not use Artificial Intelligence (AI) in the preparation of this manuscript.

## REFERENCES

1. Bekono-Nessah I, Duah-Asante KA, Poku D, et al. Whole-Eye Transplantation: How Far Are We From a Breakthrough? Ophthalmic Plast Reconstr Surg 2024;40:597–602.

2. Scarabosio A, Surico PL, Tereshenko V, et al. Whole-eye transplantation: Current challenges and future perspectives. World J Transplant 2024;14:95009.

3. Laspro M, Chaya BF, Brydges HT, et al. Technical Feasibility of Whole-eye Vascular Composite Allotransplantation: A Systematic Review. Plast Reconstr Surg Glob Open 2023;11:e4946.

4. Laspro M, Thys E, Chaya B, et al. First-in-Human Whole-Eye Transplantation: Ensuring an Ethical Approach to Surgical Innovation. Am J Bioeth 2024;24:59–73.

5. Ceradini DJ, Tran DL, Dedania VS, et al. Combined Whole Eye and Face Transplant: Microsurgical Strategy and 1-Year Clinical Course. JAMA 2024;332:1551–1558.

6. Cesaretti M, Izzo A, Pellegrino RA, et al. Cold ischemia time in liver transplantation: An overview. World J Hepatol 2024;16:883–890.

7. Peters-Sengers H, Houtzager JHE, Idu MM, et al. Impact of Cold Ischemia Time on Outcomes of Deceased Donor Kidney Transplantation: An Analysis of a National Registry. Transplant Direct 2019;5:e448.

8. Tennankore KK, Kim SJ, Alwayn IPJ, Kiberd BA. Prolonged warm ischemia time is associated with graft failure and mortality after kidney transplantation. Kidney Int 2016;89:648– 658.

9. Brennan C, Sandoval PR, Husain SA, et al. Impact of warm ischemia time on outcomes for kidneys donated after cardiac death Post-KAS. Clin Transplant 2020;34:e14040.

10. Osborne NN, Casson RJ, Wood JPM, et al. Retinal ischemia: mechanisms of damage and potential therapeutic strategies. Prog Retin Eye Res 2004;23:91–147.

11. Bermudez MA, Gonzalez F. Differential sensitivity of the On and Off visual responses to retinal ischemia. Exp Eye Res 2020;191:107906.

12. Country MW. Retinal metabolism: A comparative look at energetics in the retina. Brain Res 2017;1672:50–57.

13. Tannen JN. Paving the Way for Human Eye Transplants. InventUM 2025. Available at: https://news.med.miami.edu/paving-the-way-for-human-eye-transplants/ [Accessed September 14, 2025].

14. Lemole GM, Henn JS, Zabramski JM, Spetzler RF. Modifications to the orbitozygomatic approach. 2003. Available at: https://thejns.org/view/journals/j-neurosurg/99/5/article-p924.xml [Accessed September 14, 2025].

15. Zabramski JM, Kiriş T, Sankhla SK, et al. Orbitozygomatic craniotomy. 1998. Available at: https://thejns.org/view/journals/j-neurosurg/89/2/article-p336.xml [Accessed September 14, 2025].

16. Samarage HM, Kim WJ, Zarrin DA, et al. Endoscope-Assisted Pedicled Maxillary Artery to Middle Cerebral Artery Bypass: An Anatomic Feasibility Study. Oper Neurosurg 2023;24:209– 220.

17. Okawa H, Sampath AP, Laughlin SB, Fain GL. ATP consumption by mammalian rod photoreceptors in darkness and in light. Curr Biol 2008;18:1917–1921.

18. Palmhof M, Frank V, Rappard P, et al. From Ganglion Cell to Photoreceptor Layer: Timeline of Deterioration in a Rat Ischemia/Reperfusion Model. Front Cell Neurosci 2019;13:174.

19. Block F, Schwarz M. The b-wave of the electroretinogram as an index of retinal ischemia. Gen Pharmacol 1998;30:281–287.

20. Antonetti DA, Lin C-M, Shanmugam S, et al. Diabetes Renders Photoreceptors Susceptible to Retinal Ischemia-Reperfusion Injury. Invest Ophthalmol Vis Sci 2024;65:46.

21. Schlegel A, Mueller M, Muller X, et al. A multicenter randomized-controlled trial of hypothermic oxygenated perfusion (HOPE) for human liver grafts before transplantation. J Hepatol 2023;78:783–793.

22. Chapman WC, Barbas AS, D’Alessandro AM, et al. Normothermic Machine Perfusion of Donor Livers for Transplantation in the United States: A Randomized Controlled Trial. Ann Surg 2023;278:e912–e921.

23. Pinnelas R, Kobashigawa JA. Ex vivo normothermic perfusion in heart transplantation: a review of the TransMedics® Organ Care System. Future Cardiol 2022;18:5–15.

24. Prasad NK, Pasrija C, Talaie T, et al. Ex Vivo Lung Perfusion: Current Achievements and Future Directions. Transplantation 2021;105:979–985.

25. Coxworth B. Portable device could make functional eye transplants a reality. New Atlas 2025. Available at: https://newatlas.com/medical-devices/eye-ecma-eyeball-transplant-machine/ [Accessed September 14, 2025].

26. Anon. Scientists Create Device That Could Make World’s First Human Eye Transplant Possible | Reuters Connect. Available at: https://www.reutersconnect.com/item/scientists-create-device-that-could-make-worlds-first-human-eye-transplant-possible/dGFnOnJldXRlcnMuY29tLDIwMjU6bmV3c21sX01UMUNWTUQ1NDk5NzQzOQ?utm_source=chatgpt.com [Accessed September 14, 2025].

27. . Health NL. The World’s First Whole-Eye & Partial-Face Transplant Recipient Achieves Remarkable Recovery, with Viable Eye One Year After Landmark Surgery. Available at: https://www.prnewswire.com/news-releases/the-worlds-first-whole-eye--partial-face-transplant-recipient-achieves-remarkable-recovery-with-viable-eye-one-year-after-landmark-surgery-302242949.html [Accessed September 14, 2025].

28. Ji T, Hou K, Li C, Yu J. Imaging features of internal maxillary artery and extracranial middle meningeal artery and their relationships on head CTA. Neuroradiol J 2021;34:629–641.

29. Gofur EM, Al Khalili Y. Anatomy, Head and Neck: Internal Maxillary Arteries. In: StatPearls. Treasure Island (FL): StatPearls Publishing; 2025. Available at: http://www.ncbi.nlm.nih.gov/books/NBK542301/ [Accessed September 14, 2025].

30. Yu Z, Shi X, Brohi SR, et al. Measurement of Blood Flow in an Intracranial Artery Bypass From the Internal Maxillary Artery by Intraoperative Duplex Sonography. Journal of Ultrasound in Medicine 2017;36:439–447.

31. Zhang F, Lineaweaver WC, Buntic R, Walker R. Mechanical evaluation of anastomotic tension and patency in arteries. J Reconstr Microsurg 1996;12:121–126.

32. Wang R, Raykin J, Brewster LP, Gleason RL. A Novel Approach to Assess the In Situ Versus Ex Vivo Mechanical Behaviors of the Coronary Artery. J Biomech Eng 2017;139:0110101–0110107.

33. Yamane Y, Uchida N, Okubo S, et al. Impact of the size mismatch between saphenous vein graft and coronary artery on graft patency. Gen Thorac Cardiovasc Surg 2017;65:25–31.

34. Gokoffski KK, Washington KM, Chuck RS. Clinical and Scientific Considerations for Whole Eye Transplantation: An Ophthalmologist’s Perspective. Transl Vis Sci Technol 2025;14:13.

